# Athlete deaths during the COVID-19 vaccination campaign: contextualisation of online information

**DOI:** 10.1101/2023.02.13.23285851

**Authors:** Mathijs Binkhorst, Daniel J. Goldstein

## Abstract

**Background and aim:** Lay people and medical professionals have suggested a link between (mRNA) COVID-19 vaccination and a purported increase in sudden cardiac arrest (SCA) and death (SCD) among athletes. We aimed to compare the athlete death rate in 2021-2022 with pre-pandemic estimates and investigate the role of vaccination.

**Methods:** A comprehensive, much referenced, publicly available list of health issues, emergencies, and SCA/SCD in athletes from January 2021 to December 2022 was analysed. Demographic data, country, type of sport, vaccination status, and possible association between reported medical events and vaccination were evaluated for the complete set of athletes. The following data were specifically assessed for cases of SCD in young US athletes and compared to matched data from pre-pandemic studies: average annual SCD number, mean age, male/female ratio, sports with highest death toll, cause and scene of death, and relation to exercise. Descriptive statistics were used.

**Results:** The list contained 1653 entries. (Former) athletes, aged 5-86 years, from 99 countries, participated in 61 different sports. In multiple cases, causes of and circumstances surrounding medical events were irretrievable. Many cases involved non-cardiovascular, exercise-unrelated aetiologies. Vaccination details were scarce. In 63 (3.8%) cases, including 9 fatal events, there was a plausible association with COVID-19 vaccination. In US athletes aged 9-40 (mean 22.7) years, 166 SCD cases were identified (average 83/year), mainly in males (83%) and in football (39.8%) and basketball (16.9%). Main causes of death were non-cardiovascular exercise-unrelated (22.9%) or unknown (50.6%). Deaths primarily occurred at rest (32.5%) or under unknown circumstances (38.6%). SCD characteristics were similar to those of two pre-pandemic studies with comparable datasets.

**Conclusion:** SCD rate among young US athletes in 2021-2022 was comparable to pre-pandemic estimates. There is currently no evidence to substantiate a link between (mRNA) COVID-19 vaccination and SCD in (young) athletes.

## Introduction

Exercise is generally regarded as beneficial for cardiovascular and overall health. Physical activity can nevertheless cause sudden cardiac arrest (SCA) and death (SCD) in athletes on the pitch or shortly after cessation of exercise (‘paradox of sport’).^1-3^ Of all out-of-hospital cardiac arrests (OHCA), approximately 6% is sports-related.^1^ The incidence of exercise-related SCD among young (≤35 years) athletes is estimated around 0.6–4.4 per 100,000 person-years, with a striking male preponderance (81-97%).^1-4^ In the United States (US), sudden death is predominantly witnessed in basketball and football players, while soccer is the main culprit in Europe.^3^ SCA/SCD usually result from a critical increase in myocardial oxygen demand and/or ventricular arrythmia associated with exercise-induced catecholamine release in individuals with underlying cardiac (arrhythmogenic) vulnerabilities, with contributory roles of heat and dehydration.^3,5^ In young athletes, these underlying aetiologies comprise various structural (hypertrophic cardiomyopathy, coronary artery anomalies, arrhythmogenic right ventricular cardiomyopathy, mitral valve prolapse, aortic dissection/rupture), electrical (Wolff-Parkinson-White, Brugada, and long QT syndromes, catecholaminergic polymorphic ventricular tachycardia), and acquired (premature coronary artery disease, commotio cordis, myocarditis) cardiac abnormalities.^2,3,5-9^ In addition, sudden death may also be provoked by trauma, asthma exacerbation, heat stroke, ruptured aneurysm, and drugs.^4,7^

In light of the present study, it is noteworthy that (viral) myocarditis accounts for 3-8% of SCD in athletes according to most studies,^2,3,6,9^ although higher and lower percentages have been reported. A recent study revealed that the pre-SARS-CoV-2 pandemic occurrence of exercise-related, myocarditis-associated SCD in competitive and recreational players was very rare, with an incidence of 0.047 per 100,000 person-years (i.e. one myocarditis-associated death per 2.13 million person-years).^10^ Physical exertion increases the risk of SCD in individuals with acute myocarditis.^11^ Myocarditis can sometimes present with SCA/SCD due to ventricular tachycardia (VT), ventricular fibrillation (VF), or atrioventricular (AV) block.^11^ In myocarditis patients, the resting electrocardiogram (ECG) may be normal or show sinus tachycardia with non-specific alterations in the ST-T segment; more deleterious changes include new left bundle branch block (LBBB), QRS abnormalities, northwest axis deviation, and QTc prolongation.^11^ Athletes with myocarditis are recommended to refrain from participation in competitive sports for 3-6 months and await cardiac re-evaluation before return to play.

Lately, numerous reports have been published, mainly in the (social) media, about young athletes succumbing ‘on the pitch’, sometimes with a speculative undertone suggesting that COVID-19 vaccines were causally involved. This media attention and conjecture basically started with the cardiac arrest of Danish soccer player Christian Eriksen and continued up to the present day, vividly reignited after the recent collapse of Buffalo Bills safety Damar Hamlin. It appears that myocarditis, as a rare side effect of mRNA-based COVID-19 vaccines, particularly in male adolescents and young men, has somehow been automatically linked to a purported increase in SCA/SCD among young athletes, although mRNA COVID-19 vaccine-induced myocarditis is mostly self-limiting with apparently benign short and medium term outcomes.^12^ Several sports cardiologists and organisations, such as the National Center for Catastrophic Sport Injury Research, American College of Cardiology, and the international football association (FIFA), have declared to be unaware of a clear association between COVID-19 vaccination and athletic deaths.^13-15^

In the online blog GoodSciencing.com, anonymous contributors have compiled a comprehensive, though unselected list of largely unverified medical events, emergencies, cardiac arrests and deaths in athletes of all ages from around the globe, based on news stories.^16^ Evidently, privacy issues restricted the availability of important details in the assembled media reports, such as vaccination status, medical test results, and exact cause of death. Nevertheless, this list is probably the most extensive, publicly available collection of emergencies and fatalities in athletes in the past two years. It has not only been massively referenced in the (social) media, but also in several peer-reviewed publications,^17,18^ and most recently in a letter to the editor.^19^ Even in these formal publications, it is suggested that sports-related SCA/SCD has (sharply) increased since – and due to – the start of the COVID-19 vaccination campaign. If these suggestions can be substantiated, there is, of course, reason for concern about the safety of the (mRNA-based) COVID-19 vaccines. However, if such assertions cannot be corroborated, they may unnecessarily have a negative impact on vaccination acceptance.

We therefore sought to determine whether the number of SCD cases among athletes in 2021-2022, based on the GoodSciencing list, markedly deviated from pre-pandemic estimates, and whether there is evidence for an allegedly detrimental role of (mRNA) vaccines in this context.

## Methods

Being unaware of a published, official registry reporting SCA/SCD in (young) athletes during the SARS-CoV-2 pandemic, or specifically since initiation of COVID-19 vaccination, we used the GoodSciencing compilation as a data source. The list was accessed on January 19^th^, 2023. At that moment, ‘1653 athlete cardiac arrests or serious issues, including 1148 fatalities, since COVID injection’ were included. After obtaining a general impression of the entries and reported diagnoses, we attempted to specifically evaluate the following characteristics of the listed athletes: demographic data, country, type of sport, vaccination status, and possible association between the reported medical events and vaccination.

Media attention has mainly been directed at young athletes. Tragedies in young individuals can have a major societal impact. For these reasons, and also because 1) entries concerning US athletes appeared to predominate in the list, and 2) suitable pre-pandemic studies for comparison seemed to be available for this category, we decided to focus our analysis on SCD in young US athletes. Consequently, we created a shortlist by extracting all cases in which US athletes <40 years died from the GoodSciencing list in the period extending from January 2021 to December 2022. Google search engine (Google, Mountain View, California, USA) was used to verify the events, uncover vaccination status, and determine the cause of and circumstances surrounding death. Information was preferentially derived from online articles stating findings from autopsy reports and communications by medical examiners and coroners. In addition, statements by police departments, obituaries, and details provided by relatives were used.

In PubMed, we searched for studies on SCD in young US athletes predating the pandemic in order to compare various findings from our analysis (average annual SCD number, mean age, male/female ratio, sports with highest death toll, cause and situation of death, and whether or not cases were associated with genuine exercise-related, cardiovascular conditions) with those from earlier registries.

We used descriptive statistics to analyse the data in Excel (version 16.0.1, Microsoft, Redmond, Washington, USA). This study was exempted from ethical approval.

## Results

The complete list contained 1653 entries of health-related problems, emergencies, cardiac arrests, and fatalities in athletes from 99 countries, participating in 61 different types of sport, aged 5-86 years. Most reports (411 of 1653, 24.9%) were from the USA. Other countries with a substantial number of reports (>50) included Australia, Canada, Brazil, India, France, Spain, Germany, and Italy. Table 1 shows various conditions diagnosed in the complete set of diseased or deceased athletes. Many of these conditions were encountered several times on the list (e.g. at least 35 cancer cases) and did not represent true cardiovascular, exercise-related problems.

**Table 1.**
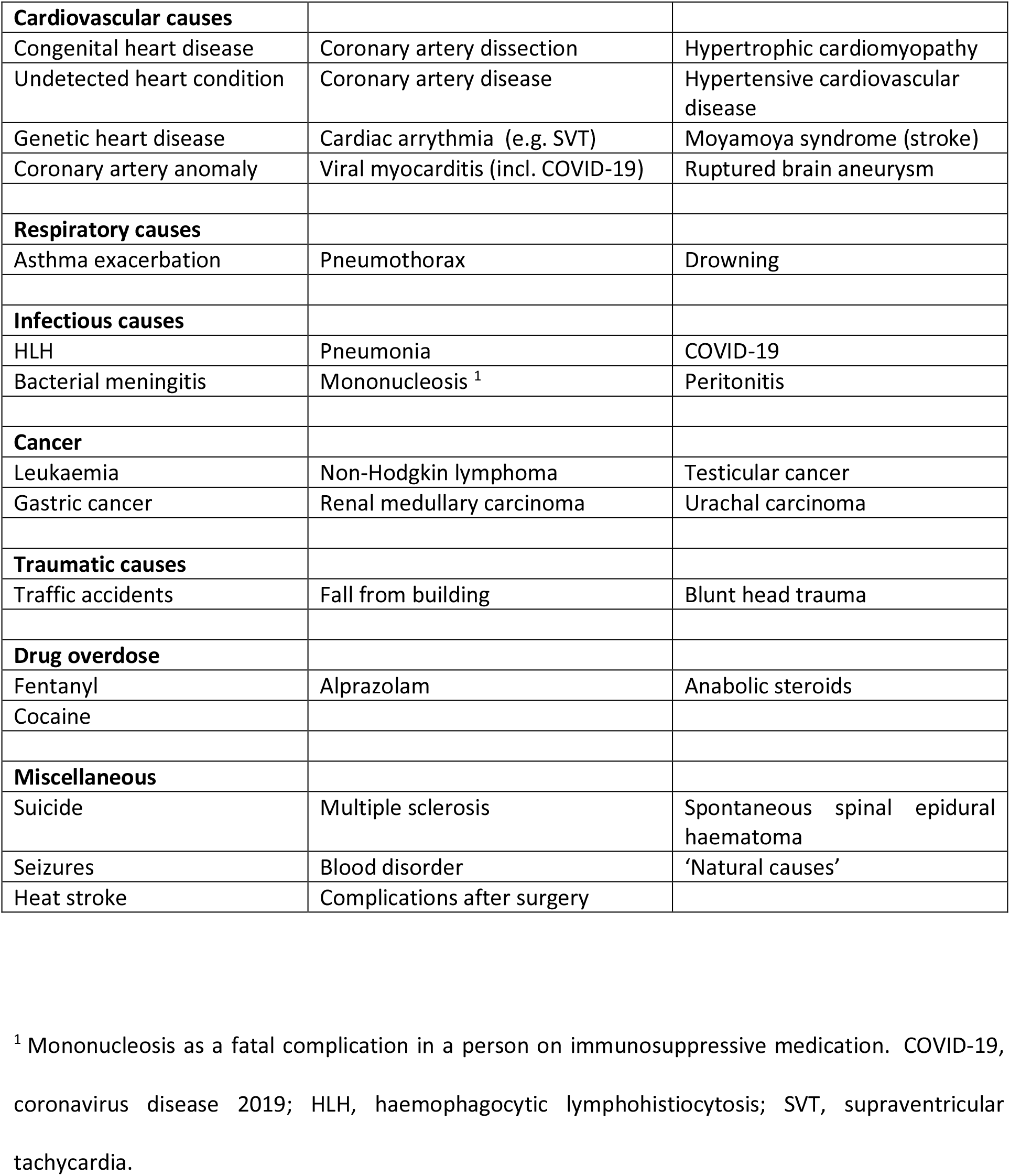
Retrievable causes of medical events and deaths in athletes of all ages worldwide

|  |  |  |
| --- | --- | --- |
| **Cardiovascular causes** |  |  |
| Congenital heart disease | Coronary artery dissection | Hypertrophic cardiomyopathy |
| Undetected heart condition | Coronary artery disease | Hypertensive cardiovascular disease |
| Genetic heart disease | Cardiac arrhythmia (e.g. SVT) | Moyamoya syndrome (stroke) |
| Coronary artery anomaly | Viral myocarditis (incl. COVID-19) | Ruptured brain aneurysm |
| **Respiratory causes** |  |  |
| Asthma exacerbation | Pneumothorax | Drowning |
| **Infectious causes** |  |  |
| HLH | Pneumonia | COVID-19 |
| Bacterial meningitis | Mononucleosis <sup>1</sup> | Peritonitis |
| **Cancer** |  |  |
| Leukaemia | Non-Hodgkin lymphoma | Testicular cancer |
| Gastric cancer | Renal medullary carcinoma | Urachal carcinoma |
| **Traumatic causes** |  |  |
| Traffic accidents | Fall from building | Blunt head trauma |
| **Drug overdose** |  |  |
| Fentanyl | Alprazolam | Anabolic steroids |
| Cocaine |  |  |
| **Miscellaneous** |  |  |
| Suicide | Multiple sclerosis | Spontaneous spinal epidural haematoma |
| Seizures | Blood disorder | 'Natural causes' |
| Heat stroke | Complications after surgery |  |

Using the (truncated) search terms *vaccin*, inject*, vax*, shot*, and *jab*, we explored the list to identify cases in which a link between vaccination and the reported medical events could be considered. In the large majority of cases, (definite) information on the athletes’ vaccination status was irretrievable; details on type, timing, and dose number of vaccinations were particularly scarce. In the full collection of 1653 reports gathered from around the globe, 124 athletes (7.5%) had a practically confirmed vaccination status and no other directly obvious cause for their disease or death. Following further scrutiny with more elaborate Google searches, 63 cases (3.8%) were deemed plausibly associated with COVID-19 vaccination (i.e. reported with sufficient detail, absence of apparent alternative cause, temporal relation to vaccination, and known side effects) (Supplementary data). Thrombotic/thrombocytopenic events occurred in some individuals after receiving adenovirus vector vaccines. Three persons developed pulmonary embolism upon vaccination with the mRNA-1273 vaccine. Myocarditis, pericarditis, and their combination were witnessed in (young) athletes vaccinated with mRNA-based vaccines. Four deaths were probably linked to the thrombotic/thrombocytopenic consequences of adenovirus vector vaccines. In 5 other cases, that appeared to be associated with vaccination (2 BNT162b2, 2 ChAdOx1, 1 type unknown), the exact cause of death was not fully elucidated. In a few cases (e.g. Christian Eriksen), relatives, coaches, club representatives, or the athletes themselves explicitly declared that the health issues were not associated with vaccination. Also, in the first months of 2021, healthy adolescents and young adults were not eligible for vaccination in most countries.

The list contained numerous unverifiable reports, in which names and/or ages were lacking. Many (fatal) events did not take place during exercise (e.g. 39 athletes died in their sleep). Multiple reports dealt with individuals >40 years (up to 86 years) old, so not ‘young’ or ‘current’ athletes. The fact that many reports did not pertain to active athletes was exemplified by the observation that the word ‘former’ appeared 194 times on the list. Moreover, a considerable number of entries did not involve athletes, but coaches (n=107), referees (n=25), linesmen (n=3), managers (n=7), and teachers (n=13) (if these persons had a dual role, e.g. coach and teacher, only one role was counted).

Looking specifically at US athletes <40 years who died between January 2021 and December 2022, we identified 166 fatal cases (average 83/year). Of these, 138 (83%) were male. Mean age was 22.7 years (range 9-40 years). Vaccine-associated myocarditis primarily affects male individuals 12-29 years of age. In our shortlist, 3 children were <12 years and 36 adults were >30 years. The young athletes participated in 33 types of sport, mainly in American football (39.8%) and basketball (16.9%).

Causes of death in the subset of young US athletic casualties are shown in Table 2. In most cases, the aetiology was either unknown (50.6%) or involved a non-cardiovascular, exercise-unrelated condition (22.9%). Deaths occurred during physical activity in 24.7%, shortly after exercise in 3.6%, and at rest in 32.5% (Table 3). One person died during surgery. In the remaining cases (38.6%), the scene of the tragic incident could not be reliably retrieved from the news sources.

**Table 2.**
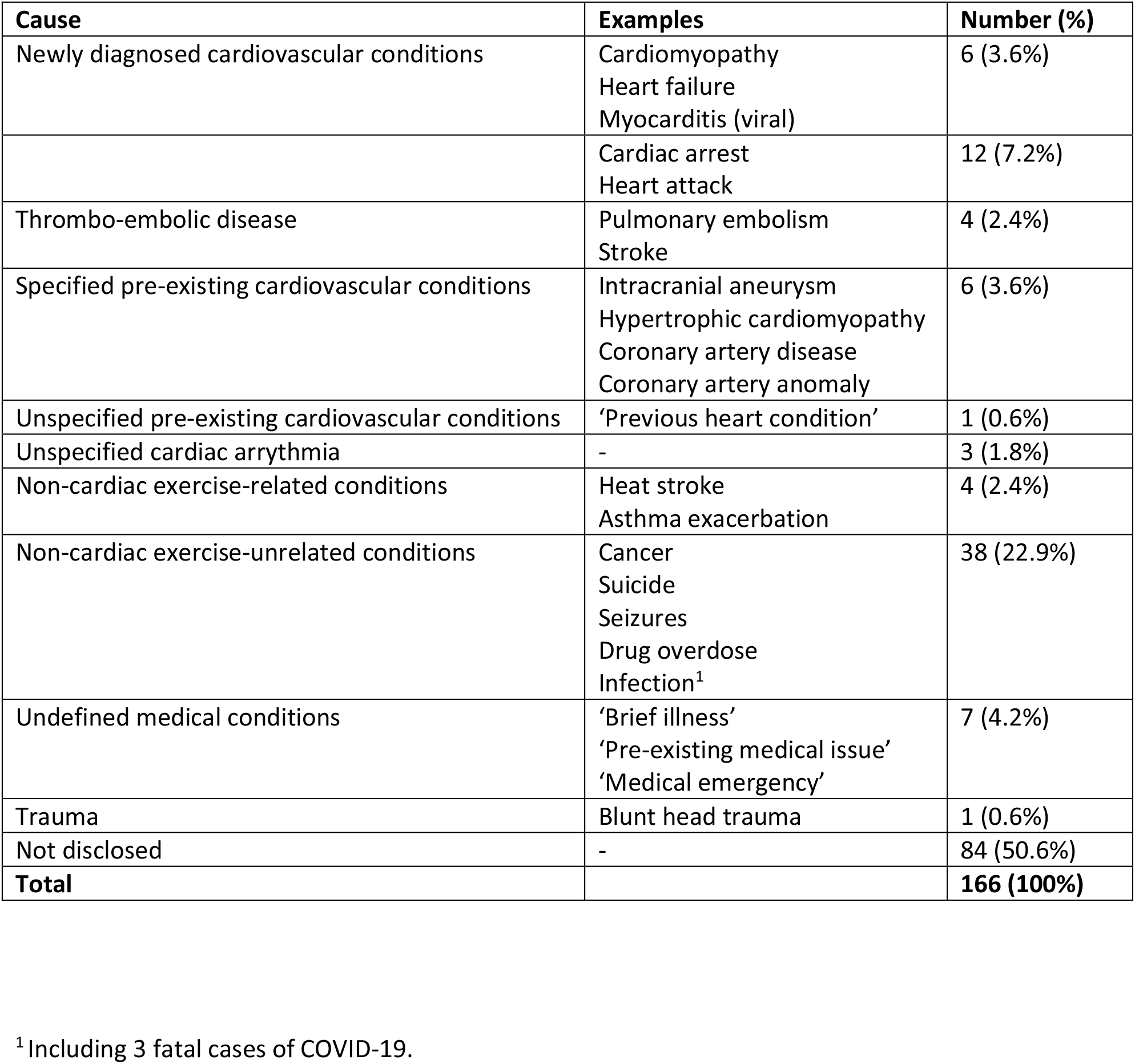
Causes of death in US athletes <40 years

| **Cause** | **Examples** | **Number (%)** |
| --- | --- | --- |
| Newly diagnosed cardiovascular conditions | Cardiomyopathy<br>Heart failure<br>Myocarditis (viral) | 6 (3.6%) |
|  | Cardiac arrest<br>Heart attack | 12 (7.2%) |
| Thrombo-embolic disease | Pulmonary embolism<br>Stroke | 4 (2.4%) |
| Specified pre-existing cardiovascular conditions | Intracranial aneurysm<br>Hypertrophic cardiomyopathy<br>Coronary artery disease<br>Coronary artery anomaly | 6 (3.6%) |
| Unspecified pre-existing cardiovascular conditions | 'Previous heart condition' | 1 (0.6%) |
| Unspecified cardiac arrhythmia | - | 3 (1.8%) |
| Non-cardiac exercise-related conditions | Heat stroke<br>Asthma exacerbation | 4 (2.4%) |
| Non-cardiac exercise-unrelated conditions | Cancer<br>Suicide<br>Seizures<br>Drug overdose<br>Infection<sup>1</sup> | 38 (22.9%) |
| Undefined medical conditions | 'Brief illness'<br>'Pre-existing medical issue'<br>'Medical emergency' | 7 (4.2%) |
| Trauma | Blunt head trauma | 1 (0.6%) |
| Not disclosed | - | 84 (50.6%) |
| **Total** |  | **166 (100%)** |

**Table 3.**
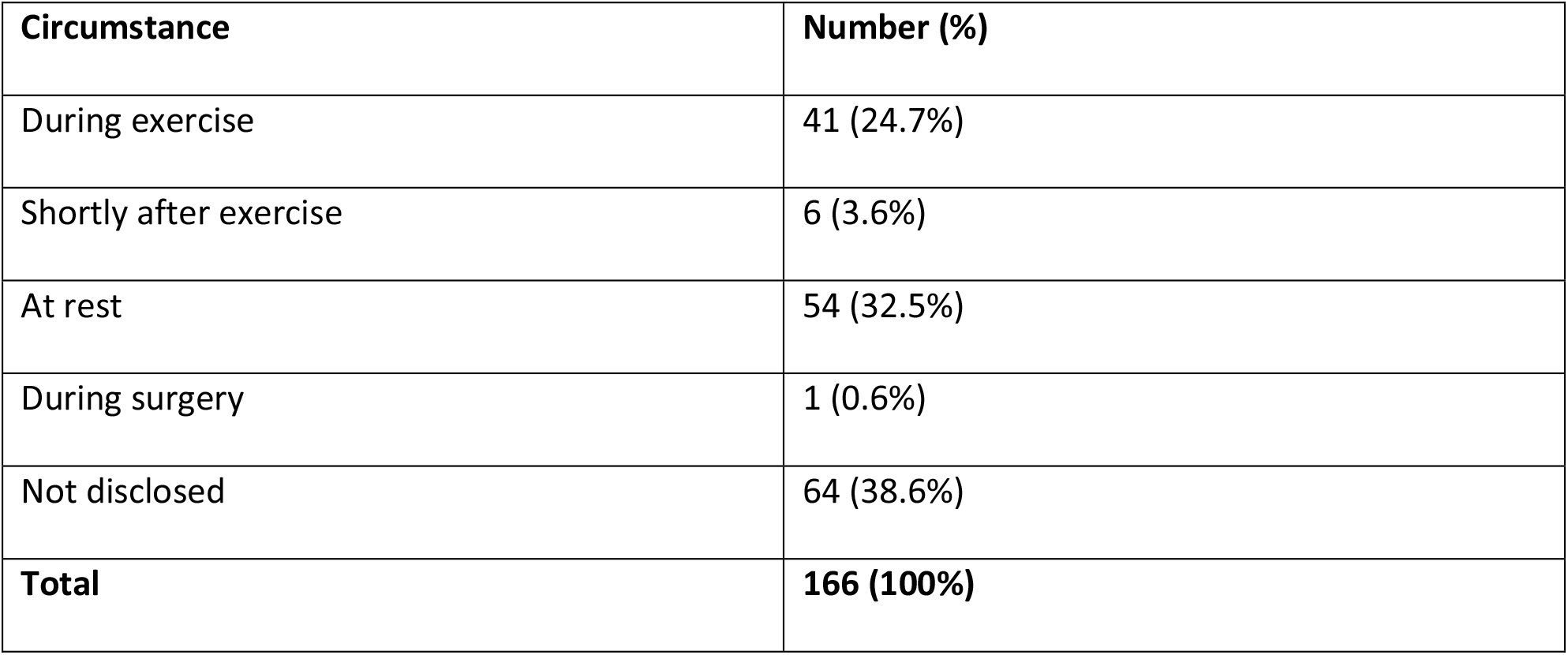
Circumstances surrounding fatal events in US athletes <40 years

Our PubMed search yielded 3 potentially useful publications for comparison with the GoodSciencing list. Of these, the study by Maron et al. was considered most suitable, since these authors included 1) similar ages (8-39 years); 2) athlete deaths only from the US; 3) a more or less comparable case mix in terms of aetiologies, although traumatic causes (25%) and probable or proven cardiovascular causes (56%) predominated in Maron’s study, while undisclosed and non-cardiac causes were most prevalent in our shortlist; 4) fatal events during or just after exercise (80%) and at rest (20%); and 5) many different types of sport (38 in a registry covering 27 years).^7^ Based on this registry (1980-2006), an average of 66 deaths per year (range 50-76) was calculated. Mean age was 19 years, 89% was male, and football (30%) and basketball (22%) were the sports with the highest number of sudden deaths in this study.

Peterson et al. assembled SCDs in (active and recently retired) US athletes from 2014-2018, which occurred either during (63.6%), within 1 hour after (3.5%), or unassociated with (27.2%) exercise.^8^ They included competitive athletes aged 11-29 years, 16 different types of sport, and predominantly reported on cardiac causes (87.5%). This study identified 173 confirmed SCD cases (and 158 SCA cases with survival), so on average 43 SCDs per year. Mean age was 16.9 years, 81.5% was male, and basketball (30%) and football (29%) were the main sports involved. When restricting our analysis to the inclusion criteria of Peterson et al. (ages 11-29 years, only cardiac and undisclosed causes), we arrived at a number of 83 deaths in two years (42/year). In our adjusted analysis, mean age was 18.8 years, 83.1% was male, and cause of death was cardiac in 24.1% and undisclosed in 75.9%.

Findings from the study by Bille et al. were considered insufficiently appropriate for comparison with the data in our shortlist.^9^ In that study, causes of SCD in athletes ≤35 years participating in (at least) 25 different sports were systematically reviewed. However, the study focused on strictly exercise-related events with true cardiac causes. Also, these authors did not specify from which (number of) countries their data were derived (from the references it can be concluded that multiple countries were included, e.g. US, Italy, Israel, Sweden, Spain, and France).

## Discussion

We performed a thorough analysis of a comprehensive, publicly available, and much referenced collection of health-related problems, medical emergencies, sudden cardiac arrests and deaths in athletes, which occurred over the past two SARS-CoV-2 pandemic years and since the initiation of the COVID-19 vaccination campaign. We mainly found that the SCD rate among young US athletes in 2021-2022 was in the same range as pre-pandemic estimates. Our analysis revealed a slightly higher average annual SCD number (83 per year) than Maron et al. (66 (range 50-76) per year).^7^ However, it should be noted that the latter study mainly included systematically tracked and rigorously verified cases of actual exercise-related sudden death with established traumatic and cardiovascular aetiologies, whereas our dataset contained considerably less verifiable cases, with many undisclosed, non-cardiac, and exercise-unrelated causes of death. Our findings neatly matched those reported by Peterson et al.^8^ Again, these authors predominantly documented confirmed cases of sports-related fatalities due to cardiovascular pathologies, contrary to the many unknowns encountered while analysing the GoodSciencing list.^16^ Other aspects of the studied cases of SCD in young athletes, such as the male/female ratio and sports with the highest death toll, also mirrored the findings from pre-pandemic publications.^3,7,8^

Based on our analysis, evidence appears to be lacking to link COVID-19 vaccines to (a purported increase in) SCD in (young) athletes. In the total list of 1653 entries, we identified 63 cases with a plausible relation to COVID-19 vaccination. These mostly concerned recognised, rare side effects of the adenovirus vector (thrombosis/thrombocytopenia) and mRNA-based (myocarditis/pericarditis) vaccines. Four deaths – essentially exercise-unrelated – were likely associated with the just mentioned coagulation disturbances sporadically observed after administration of adenovirus vector vaccines. Four out of five additional fatalities were temporally closely related to vaccination, but causality could not be established. While scientifically sound studies on the correlation between COVID-19 vaccination on the one hand and cardiac arrest and death on the other are scarce for the general population, they are virtually nonexistent for athletes. Nevertheless, a recent Italian cohort study demonstrated that vaccinated individuals had a significantly lower all-cause mortality and reduced risk of stroke, thrombo-embolism, and cardiac arrest compared to unvaccinated controls.^20^

Although myocarditis is a generally acknowledged, rare side effect of mRNA-based COVID-19 vaccines, and can be complicated by ventricular arrhythmia and SCA/SCD, it is a rather infrequent cause of SCD,^2,3,6,9,10^ also in the present study (Tables 1 and 2), and there is currently no evidence to attribute significant numbers of young athlete deaths to this vaccine-induced condition. Male adolescents and male young adults (12-29 years) are mainly affected by myocarditis (incidence 1 in 6,800–35,000), usually 2-4 days after the second mRNA dose.^21-23^ In general, the risk of cardiac complications, including myocarditis, is much higher following SARS-CoV-2 infection than after mRNA vaccination.^21^ According to Patone et al., vaccination also provided some protection against the cardiovascular consequences of COVID-19.^21^ However, in the subgroup of young males, the risk of myocarditis after a second dose of the mRNA-1273 vaccine was higher than after SARS-CoV-2 infection.^21,22^ Despite the current absence of an established relationship between mRNA vaccination and (increasing) athlete deaths, the risk-benefit ratio of mRNA(−1273) vaccination must be carefully considered in this subgroup nowadays, since multisystem inflammatory syndrome in children (MIS-C) has become less of a threat, the virus has evolved into a less virulent (Omicron) variant, and these young individuals have already attained a considerable level of (hybrid) immunity.

The short and medium term outcomes of mRNA vaccine-associated myocarditis were apparently favourable in a recent follow-up surveillance study.^12^ Eighty-one percent of US adolescents and young adults (12-29 years) was clinically recovered at least 90 days after myocarditis onset, with improvement or normalisation of the electrocardiogram (ECG), echocardiogram, troponin levels, ambulatory rhythm monitoring, and exercise stress testing in the large majority; 68% could safely return to play. No deaths were reported in this follow-up study. Absence of haemodynamic instability, ECG normalisation, and preserved left ventricular systolic function on the echocardiogram are indeed associated with a good prognosis.^11^ However, the presence of late gadolinium enhancement (LGE) on cardiac magnetic resonance imaging (MRI) – witnessed in 47% of post-vaccination myocarditis patients with a follow-up cardiac MRI – adds some uncertainty to the prognosis, since myocardial fibrosis can act as an arrhythmogenic substrate and cause ventricular arrythmia and SCA/SCD.^11^ More research is needed to determine the longer term evolution of residual LGE in patients with vaccine-induced myocarditis and the exact role of cardiac MRI in prognostication and return-to-play decisions.^11,12^

Clinically *manifest* myocarditis is often diligently monitored by cardiologists. Athletes diagnosed with myocarditis are advised to abstain from strenuous exercise for 3-6-months and should be re-evaluated for normalisation of test results before return to play.^3,11^ The incidence and significance of *subclinical* myocarditis are uncertain. Cardiac pathology is probably mild and prognosis is expected to be good in subclinical myocarditis,^11^ but the question remains – in particular for those considering a link between COVID-19 vaccination and athletic casualties – how often undetected myocarditis constitutes the underlying aetiology of SCA/SCD. Although it is currently impossible to conclusively answer this question, a hint to an answer may be found in the literature on COVID-19 itself. That is, 1.8% of US athletes had subclinical myocarditis upon screening after SARS-CoV-2 infection, while exercise-related cardiac events clearly associated with COVID-19 have been hardly documented in athletes.^24,25^

Apart from myocarditis, other cardiovascular and thrombo-embolic problems (pulmonary embolism, stroke, acute myocardial infarction) have not been associated with (monovalent) mRNA COVID-19 vaccines,^26,27^ and if so (in the case of venous and arterial thrombosis), the risk-benefit ratio has been clearly in favour of mRNA-based vaccination, for the cardiovascular and haematological risks of SARS-CoV-2 infection were considerably more pronounced than those of mRNA vaccines.^28^ Patone et al. showed that cardiac arrhythmia was not related to BNT162b2 vaccination, and only minimally linked to a second dose of the mRNA-1273 vaccine.^22^ Again, the risk of arrythmia was much higher following SARS-CoV-2 infection. Also, ventricular arrhythmias were infrequent in this study (VT 2.1%, VF 0.7%). In Taiwan, researchers conducted an interesting study, in which they compared ECGs before and after the second BNT162b2 dose in 4928 senior high school students (12-18 years).^29^ They found significant post-vaccination ECG changes in 51 (1%) students, mostly asymptomatic. Alterations included ST-T changes, premature ventricular contractions (PVC), sinus bradycardia, atrial tachycardia, incomplete right bundle branch block, QRS axis change, and mild QT prolongation. In only one male student, subclinical mild myocarditis was suspected due to PVC and slightly elevated cardiac troponin T. Four additional male teenagers had significant arrhythmia (sinus bradycardia, atrial tachycardia, PVC) according to paediatric cardiologists. Heart rates/rhythms and cardiac test results normalised in these 5 teenagers and 1 month later no adolescents were diagnosed with myocarditis or other serious adverse cardiac events. All symptoms resolved spontaneously and there were no hospitalisations. In the overall study population, there was a statistically significant, but clinically irrelevant increase in heart rate (2.6 beats/min) and decrease in QRS duration (1.1 ms) and QTc interval (2.1 ms) after vaccination. The authors concluded that the incidence of clinical and even subclinical myocarditis was low in BNT162b2 vaccine recipients, and that the BNT162b2 vaccine is unlikely to cause life-threatening arrhythmia in healthy young individuals.^29^

Hopefully, lay people and medical professionals will rely on verified and properly analysed data to arrive at substantiated conclusions regarding vaccination effects in the years to come. Himelfarb and colleagues have, among others, delineated the consequences of COVID-19 misinformation.^30^ Misleading or false assertions probably led to vaccine skepticism and refusal in 2.3 million Canadians. If these vaccinations had been administered, approximately 198,000 fewer cases, 13,000 fewer hospitalisations, and 2,800 fewer COVID-19-related deaths would have occurred in Canada, and $300 million in health-related costs could have been saved.

The main strength of our study lies in the careful matching of our data with those of suitable pre-pandemic registries, which enabled us to make a useful comparison. Our study also had some limitations. Underreporting probably occurred on both sides of the equation, that is in the GoodSciencing list,^16^ but also in the studies used for comparison.^7,8^ The data on GoodSciencing.com are not compiled in a spreadsheet, but as non-uniform text reports, which made it impractical to assemble, arrange, and analyse the data. Many causes of and circumstances surrounding death and vaccination details could not be retrieved. Due to a lack of time, finances, and resources, we were not able to verify cases in a similarly meticulous fashion as Maron and Peterson had done;^7,8^ we only used online searches to collect and verify information. However, in anticipation of the publication of more formal registries of SCA/SCD in athletes during the pandemic, we endeavoured to show that online information, such as presented on GoodSciencing.com, can be contextualised and unsubstantiated conclusions can be tackled by carefully studying publicly available news sources. Lastly, we mainly focused on one particular age group from one country. Therefore, our findings may not be generalisable to other populations.

## Conclusion

The number of SCDs among young US athletes in 2021-2022 was comparable to pre-pandemic estimates. A link between (mRNA) COVID-19 vaccination and SCD in (young) athletes cannot be substantiated with currently available evidence.

## Data Availability

The data, on which this analysis was based, are publicly available at the GoodSciencing website: https://goodsciencing.com/covid/athletes-suffer-cardiac-arrest-die-after-covid-shot/. The excerpt of the data from the online list, created by the authors in Microsoft Excel and used for this manuscript, can be obtained from the authors on reasonable request.

