## Supplementary data for "Athlete deaths during the COVID-19 vaccination campaign: contextualisation of online information"

**Supplementary data.** Cases with plausible relation to COVID-19 vaccination

| **No.** | **Adverse event** | **Vaccine type** | **Dose no.** | **Time since vaccine** | **Comments** |
| --- | --- | --- | --- | --- | --- |
| 1 | Auto-immune reaction | Unknown | 2 | Few days |  |
| 2 | Pericarditis | Unknown | 3 | Unknown |  |
| 3 | Pulmonary embolism | mRNA | Unknown | Same day | APS diagnosis |
| 4 | Dizziness/headache | BNT162b2 | 1 | Unknown |  |
| 5 | Myocarditis | Unknown | Unknown | Unknown |  |
| 6 | Myocarditis | Unknown | Unknown | Unknown |  |
| 7 | Tachycardia, fatigue,  sweating, dizziness | Unknown | 1 and 2 | Unknown |  |
| 8* | Thrombosis | ChAdOx1 | Unknown | Unknown | VITT/CVST (?) |
| 9 | Pericarditis | BNT162b2 | Unknown | Unknown |  |
| 10 | Pulmonary embolism | mRNA-1273 | 2 | Unknown |  |
| 11 | Bell’s palsy | Unknown | Unknown | Unknown |  |
| 12 | Pericardial injury | mRNA-1273 | Unknown | Unknown |  |
| 13 | Neurological, vascular | mRNA-1273 | Unknown | Unknown |  |
| 14 | Pericarditis | Unknown | 2 | Unknown |  |
| 15 | Pericarditis | Unknown | Booster | 5 days |  |
| 16 | Myocarditis, POTS | mRNA-1273 | 1 | 1 month |  |
| 17 | Myocarditis | Unknown | 2 | 12 hours |  |
| 18 | Pericarditis | BNT162b2 | Booster | 5 days |  |
| 19 | Seizures | Unknown | 2 | Unknown |  |
| 20 | Jaundice, pericarditis | Unknown | Unknown | Unknown |  |
| 21 | Myocarditis | Unknown | 3 | Unknown |  |
| 22 | Pericarditis | BNT162b2 | Unknown | Unknown |  |
| 23 | Myocarditis | Unknown | booster | Unknown | Recovered |
| 24 | Pericarditis | BNT162b2 | 2 | 5 days |  |
| 25 | Migraine, confusion,  speech difficulty,  paralysis LE | Unknown | 2 | Unknown |  |
| 26 | Pericarditis, paralysis LE | BNT162b2 | Unknown | Unknown |  |
| 27 | Myopericarditis | Unknown | Unknown | Unknown | Recovered |
| 28* | SCD | BNT162b2 | Unknown | 3 days | CoD unclear |
| 29 | Myocarditis | Unknown | 1 | 2 days |  |
| 30 | Thrombo-embolism | BNT162b2 | Unknown | Unknown |  |
| 31* | ITP | Unknown | 2 | Unknown | CoD ITP |
| 32 | Tachycardia | Unknown | Unknown | Unknown |  |
| 33 | Pericarditis | BNT162b2 | Unknown | Unknown |  |
| 34 | Myocarditis | Unknown | Unknown | Unknown |  |
| 35 | Paralysis | Ad26.COV2.S | Unknown | Unknown |  |
| 36 | Pulmonary embolism | mRNA-1273 | Unknown | Unknown |  |
| 37 | Myocarditis | Unknown | 2 | 4 days |  |
| 38 | Pulmonary embolism | mRNA-1273 | Unknown | Unknown |  |
| 39 | Moderate heart failure | Unknown | 2 | Unknown |  |
| 40* | Seizures, cardiac arrest | Unknown | Unknown | Few hours | CoD unclear ^1^ |
| 41 | Pericarditis | BNT162b2 | 2 | Unknown |  |
| 42 | Myocarditis | BNT162b2 | Unknown | Unknown | Recovered |
| 43 | Myocarditis | BNT162b2 | Unknown | Unknown | Recovered |
| 44 | Paralysing pains | BNT162b2 | Unknown | Unknown |  |
| 45* | Death | Ad26.COV2.S | Unknown | Few days | VITT (?) |
| 46* | Acute heart failure | ChAdOx1 | Unknown | 1 day | CoD unclear |
| 47 | Myocarditis | BNT162b2 | Unknown | Unknown |  |
| 48 | Myocarditis | BNT162b2 | Unknown | Unknown |  |
| 49 | Pericarditis | BNT162b2 | Unknown | Unknown |  |
| 50 | Myopericarditis | mRNA-1273 | Unknown | Unknown |  |
| 51* | Death | BNT162b2 | Unknown | 11 days | CoD unclear |
| 52* | Cerebral haemorrhage,  thrombocytopenia | ChAdOx1 | Unknown | 11 days | VITT/ITP (?) |
| 53 | Thrombosis | ChAdOx1 | Unknown | Unknown |  |
| 54 | Myocarditis | BNT162b2 | 2 | Unknown |  |
| 55 | Thrombosis | Ad26.COV2.S | Unknown | Unknown |  |
| 56 | Debilitating pains | BNT162b2 | 2 | Unknown |  |
| 57* | Death | ChAdOx1 | Unknown | 3 days | CoD unclear |
| 58 | Thrombosis | BNT162b2 | 1 | Unknown |  |
| 59 | Pericarditis | Unknown | 2 | Unknown |  |
| 60 | Stroke | BNT162b2 | Unknown | Unknown |  |
| 61 | Myopericarditis | Unknown | 2 | Unknown |  |
| 62 | Functional neurological  disorder | Unknown | 2 | Unknown |  |
| 63 | Cardiac arrest | BNT162b2 | 1 | 6 days |  |

**Legend to Table with supplementary data**

Cases with an asterisk (*) were fatal. ^1^ Death certificate mentioned acute coronary insufficiency, cardiopulmonary insufficiency, and pulmonary oedema. APS, antiphospholipid syndrome; CoD, cause of death; CVST, cerebral venous sinus thrombosis; ITP, immune thrombocytopenia; LE, lower extremity; POTS, postural orthostatic tachycardia syndrome; SCD, sudden cardiac death; VITT, vaccine-induced immune thrombotic thrombocytopenia.
